# Push-off in human walking emerges from support-limited feasibility not propulsion capacity

**DOI:** 10.64898/2026.05.15.26353313

**Authors:** Seyed-Saleh Hosseini-Yazdi, Karson Fitzsimons, John EA Bertram

## Abstract

Late stance push-off is widely interpreted as the mechanical expression of propulsion capacity in human walking. Here we show that this interpretation is incomplete: push-off is not a direct consequence of forward propulsion, but a conditional outcome governed by system level mechanical feasibility. Using an analytical model of step-to-step transition with empirical measurements of post-stroke hemiparetic walking, we identify two feasibility boundaries that constrain push-off. The first is a support threshold, defined by the minimum vertical force required to maintain body weight support during double support. The second is a higher transition requirement associated with redirecting the center of mass (COM) between successive stance limbs. Optimization of force limited transitions predicts that late stance push-off is mechanically infeasible below the support threshold, emerges abruptly once support feasibility is satisfied, and increases progressively toward transition defined work as available force capacity increases. Empirical analyses of ground reaction force derived COM power confirm these predictions. Forward directed propulsive impulse persists even when push-off work is absent, demonstrating that propulsion can occur without performing positive COM work. Push-off emerges only when system-level vertical feasibility becomes sufficient, and its subsequent growth is associated with increased vertical unloading and redistribution of mechanical work across the gait cycle. These results establish a hierarchical organization of walking mechanics in which feasibility precedes efficiency. Vertical support feasibility governs push-off emergence, whereas the transition requirement governs its mechanically sufficient expression. The abrupt loss of push-off therefore reflects a change in mechanically feasible locomotor solutions rather than reduced propulsive capacity.

## Introduction

Human walking is a paradigmatic example of how coordinated neuromuscular control and whole-body mechanics interact to produce efficient movement (Cavagna et al. 1977; Soo and Donelan 2010). At its core, a stride is governed by the redirection of the body’s center of mass (COM) between successive steps, a process achieved through the step-to-step transition in which mechanical energy is dissipated at heel strike and restored through push-off from the trailing limb (J. Maxwell Donelan et al. 2002; Kuo 2002; Hosseini-Yazdi and J.E.A. Bertram 2025). This transition is a primary determinant of walking energetics, with its timing and magnitude shaping both stability and energetic efficiency (Kuo 2002; Adamczyk and Kuo 2009; Hosseini-Yazdi and Bertram 2025 Sept 6). Yet despite extensive characterization of these dynamics, the fundamental conditions under which push-off emerges—and fails—remain unresolved (Sawicki and Ferris 2009; Hosseini-Yazdi and J.E. Bertram 2025).

Post-stroke hemiparetic walking provides a critical testbed for identifying these conditions. A defining feature of post-stroke hemiparetic gait is the reduction or complete absence of late-stance push-off (Hosseini-Yazdi et al. 2026b; Hosseini-Yazdi et al. 2026a), accompanied by slower walking speeds, elevated metabolic cost, and pronounced interlimb asymmetry (Olney and Richards 1996; Balasubramanian et al. 2007; White et al. 2007; Hall et al. 2011; Farris et al. 2015). These deficits are widely attributed to impaired plantar flexor force generation or disrupted neuromuscular coordination, implying that propulsion scales continuously with muscle capacity (Beaman et al. 2010; Sheffler and Chae 2015). However, this interpretation cannot account for a consistent empirical observation: at sufficiently low walking speeds or levels of impairment, late-stance push-off disappears abruptly rather than diminishing gradually (Hosseini-Yazdi et al. 2026b).

This abrupt loss indicates that push-off is not governed solely by local muscle capacity, but by global mechanical constraints acting on the whole body. Forward progression is inseparably coupled to vertical load bearing(Bertram 2005), as the COM must be supported against gravity (Dar et al. 2023) while being redirected between limbs (Adamczyk and Kuo 2009). When vertical support is insufficient, stance duration shortens (Jung and Kim 2015), limb extension is limited, and the temporal window required for push-off collapses (Neptune et al. 2001; Anderson and Pandy 2003; Liu et al. 2008). Under these conditions, push-off is not merely reduced—it must become mechanically unavailable as a controllable output.

Here we show that late-stance push-off is governed by a feasibility constraint imposed by whole-body mechanics. Specifically, a minimum level of vertical support capacity is required to maintain the COM combined vertical and horizontal trajectory during double support. Below this threshold, the step-to-step transition reorganizes such that net positive work during late stance cannot be generated. The disappearance of push-off therefore reflects a **mechanical bifurcation**: a transition between push-off infeasible and push-off capable regimes arising from continuous variation in support capacity. This implies that the absence of push-off does not necessarily reflect reduced effort, but a fundamental loss of mechanically feasible solutions for step-to-step transition.

Despite longstanding recognition of the energetic importance of step-to-step transitions, no quantitative framework has linked vertical load-bearing capacity to the emergence of push-off. It remains unclear whether a critical support threshold governs the onset of late-stance positive work, and how such a threshold manifests in empirically measurable COM energetics and interlimb coordination (Allen et al. 2011; Alam et al. 2022). Resolving this distinction is essential for determining whether propulsion deficits arise from impaired force generation or from more fundamental constraints on mechanical feasibility. That is, is hemiparetic gait a failure to emulate normal walking, or a successful attempt to provide mobility under the applied mechanical constraints?

To address this, we combine analytical modeling of step-to-step transition feasibility with empirical measurements of COM mechanical power derived from ground reaction forces in individuals with post-stroke hemiparesis walking across a range of speeds. The analytical framework identifies dual feasibility boundaries associated with vertical support and transition requirements, arising from general constraints on COM dynamics rather than population-specific impairments. Empirical analysis quantifies how system-level support capacity, limb unloading, and propulsion metrics evolve with walking speed, enabling direct evaluation of feasibility conditions governing push-off emergence.

We show that push-off is absent when vertical support capacity is insufficient to sustain COM redirection and emerges abruptly once a critical feasibility threshold is exceeded. This transition is accompanied by a reorganization of mechanical work across the gait cycle, including reduced reliance on early-stance recovery and stabilization of step-to-step dynamics. Together, these findings redefine propulsion deficits in hemiparetic walking as a consequence of system-level feasibility constraints rather than reduced propulsive capacity. More broadly, they establish that human walking operates within a constrained dynamical regime (Bertram 2005) in which key features of locomotion, including push-off, arise as emergent properties of whole-body mechanics.

## Materials and Methods

### Analytical framework: feasibility constraints in step-to-step transition

#### Step geometry and force representation

Walking was modeled using a reduced-order step-to-step transition framework in which the body’s center of mass (COM) is redirected between successive stance limbs (Alexander 1992; Alexander 1995). Step length was prescribed using a speed-dependent empirical relationship (Kuo 2001):

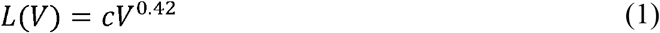

where *V* is walking speed and *c* is a proportionality constant. Assuming symmetric steps and normalized leg length, the stance limb angle relative to vertical was defined by step geometry as (Kuo 2002):

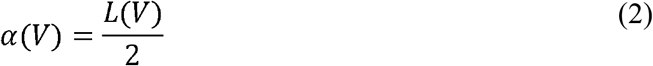

Ground reaction force was assumed to act along the stance limb and was decomposed into vertical and anterior-posterior components:

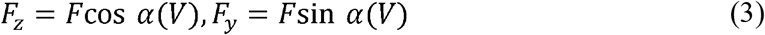

where *F* is the resultant leg force.

### COM mechanical power and push-off work

Instantaneous COM mechanical power was defined as the dot product of ground reaction force and COM velocity:

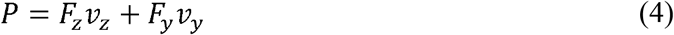

Late-stance push-off work was defined as the positive COM work generated during the double-support interval from contralateral heel strike to ipsilateral toe-off:

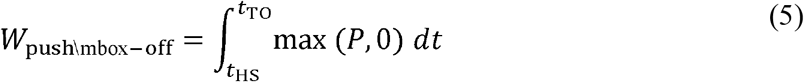

This definition isolates the mechanically relevant contribution of the trailing limb to the step-to-step transition.

### Feasibility constraints governing push-off

Push-off generation was evaluated under two feasibility conditions arising from whole-body mechanics.

### Vertical support feasibility

To maintain body-weight support during stance, the vertical force must satisfy:

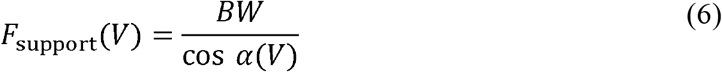

where *BW* is body weight. This defines the minimum force required to sustain COM support against gravity (Fig. 1a).

**Fig. 1:**
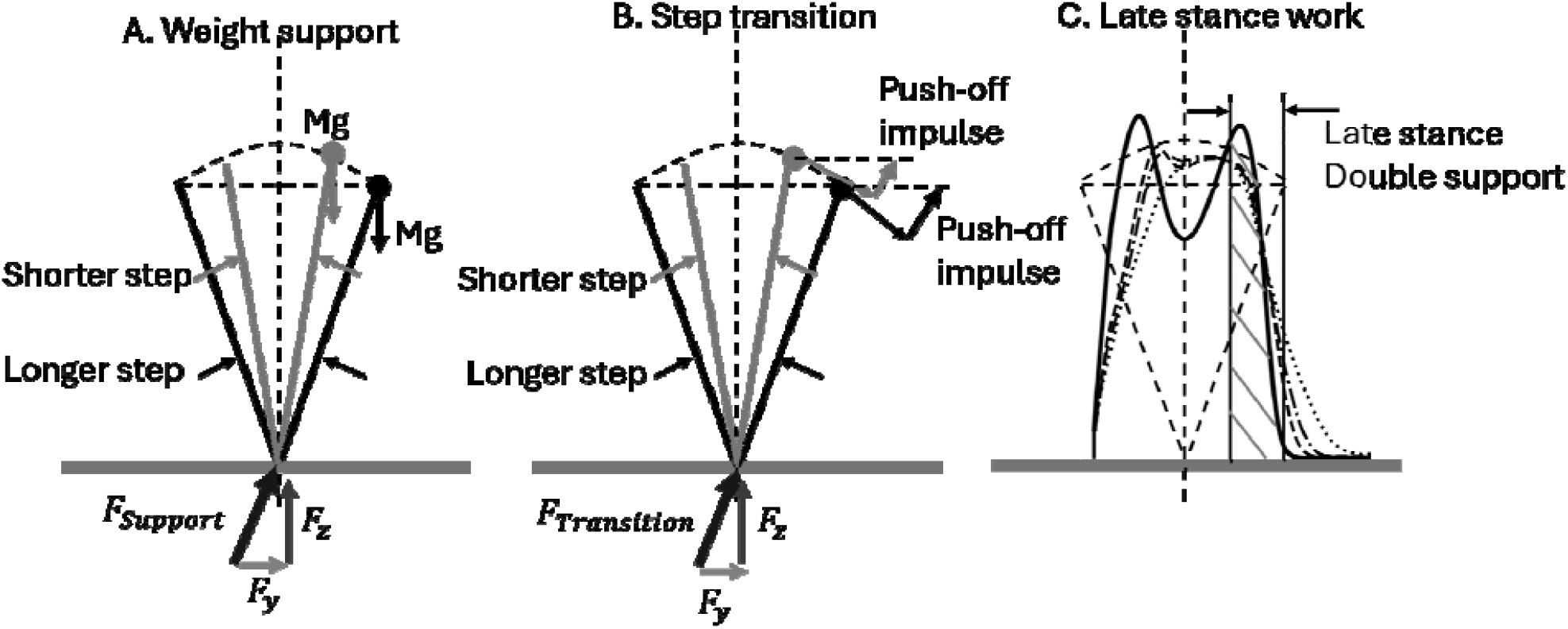
A feasibility-based interpretation of push-off in walking. Schematic illustrating the distinction between propulsion and push-off. Propulsion reflects anterior–posterior force generation, whereas push-off corresponds to positive center-of-mass (COM) work during late stance. The framework proposes two feasibility boundaries: a support threshold required for vertical loa bearing (a) and a higher transition requirement associated with COM redirection (b). Push-off emerges only once support feasibility is satisfied and increases toward transition-defined optimal work. The push-off performance interval is illustrated (c) adopted from *(Adamczyk and Kuo 2009)*.

### Transition feasibility

Push-off requirements were derived from a simplified step-to-step transition framework in which the center of mass (COM) is redirected between successive stance limbs. For a walking speed and step angle *α*(*V*), the impulse required for COM redirection was approximated as *J*_po_ = *V*tan *α* (Kuo 2002), consistent with the geometric change in COM velocity direction across steps. The corresponding push-off work was defined as 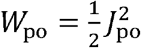 (with mass normalized) (Darici et al. 2018), reflecting the kinetic energy required for redirection. To determine feasibility, the required impulse was related to stance force capacity through a transition force threshold (Fig. 1b):

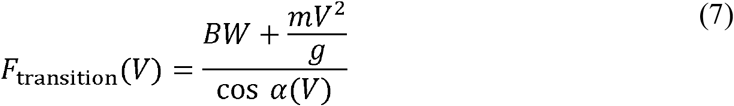

where *m* is body mass and *g* is gravitational acceleration. *F*_transition_(*V*)represents the minimum force magnitude necessary to simultaneously support body weight and generate the impulse required for COM redirection within the available stance duration. Accordingly, the transition threshold defines the force capacity required to realize the impulse *J*_po_, from which push-off work *W*_po_ follows.

Together, these conditions define distinct feasibility regimes. When available force satisfies support requirements but not transition requirements, walking remains mechanically viable, but late-stance positive work cannot be generated. This defines a push-off infeasible regime despite preserved load-bearing capacity.

### Optimization of push-off work

Push-off generation was formulated as an optimization problem in which the time-varying leg force *F* (*t*) maximizes total COM work:

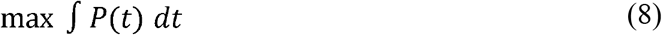

subject to the following constraints (force capacity constraint).

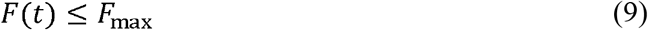

where *F*_max_ is the available force capacity.

### Vertical impulse constraint

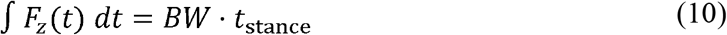

where *t*_stance_ is stance duration. This constraint enforces net vertical force balance over stance under steady-state walking conditions. This formulation determines both the existence of feasible push-off solutions and the maximum attainable work under force-limited conditions.

### Participants

Eleven individuals with post-stroke hemiparesis (5 female, 6 male; age 54.6±11.5years; time post-stroke 10.0±7.0weeks) were recruited from an inpatient rehabilitation program. Inclusion criteria required a first ischemic or hemorrhagic stroke within six months. Exclusion criteria included additional neurological conditions, significant orthopedic limitations, or comorbidities affecting walking.

All participants provided written informed consent. Empirical procedures were approved by the Conjoint Health Research Ethics Board at the University of Calgary (REB21-1576).

### Data acquisition protocol

Participants walked on a split-belt instrumented treadmill (Bertec, USA), while ground reaction forces were recorded at 1000 Hz. A safety harness was used without body-weight support, and participants were permitted to use a front handrail if required. A virtual reality environment provided visual flow consistent with treadmill speed.

Walking speed was prescribed by a supervising physiotherapist and progressed across sessions according to participant capability. Each session included at least two minutes of steady-state walking.

### COM dynamics and mechanical work

Ground reaction forces were low-pass filtered using a third-order Butterworth filter with a 10 Hz cutoff. COM acceleration was obtained by normalizing forces to body mass and integrating over time. To reduce integration drift, velocity signals were high-pass filtered at 0.256 Hz (Hosseini-Yazdi and Kuo 2025 Jan 30).

Instantaneous COM power was computed separately for each limb as:

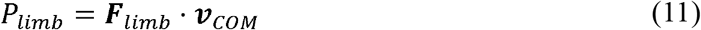

positive and negative mechanical work were obtained by time integration over the gait cycle.

### Gait event detection

Heel-strike and toe-off events were identified from vertical ground reaction force using a threshold-based swing detection method (Hansen et al. 2002; Zeni et al. 2008). Double-support intervals were defined asymmetrically for each limb to capture the mechanically relevant push-off window (Fig. 1c).

### Propulsion and support metrics

#### Propulsive Capacity Index (PCI)

Propulsive impulse was computed from the anterior-posterior ground reaction force during late stance (late double support: late_DS):

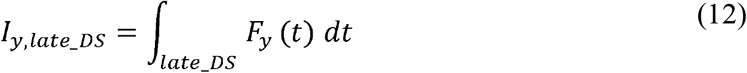

The Propulsive Capacity Index (PCI) was defined as a normalized (by vertical impulse) measure of forward-directed impulse, providing a limb-level measure of propulsion that may remain positive even when system-level feasibility for push-off is lost.

#### System vertical capacity

System-level vertical capacity was quantified from the combined vertical forces of both limbs during late double support:

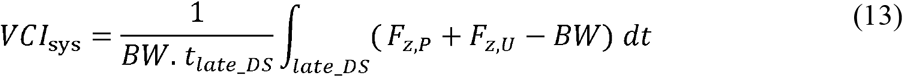

Push-off was considered feasible only when *VCI*_sys_ exceeded a minimal threshold and forward propulsion was present.

#### Vertical unloading impulse

Vertical unloading impulse was defined as:

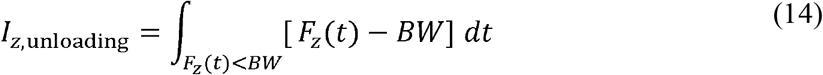

computed over stance intervals in which vertical force fell below body weight. This metric reflects interlimb load transfer during transition.

### Recovery and collapse metrics

#### Post-heel-strike recovery index (PRI)

Early-stance recovery was quantified as positive COM work following heel strike:

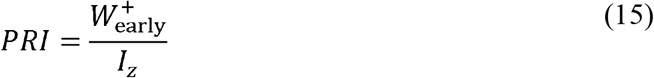

Where 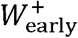 is the positive COM work during early double support.

#### Recovery Demand Support Fraction (RDSF)

The Recovery Demand Support Fraction (RDSF) quantifies available vertical support relative to body weight during early stance, capturing the mechanical feasibility of recovery following collision:

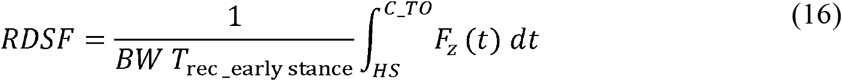

#### Limb support share

To quantify how vertical support is distributed between limbs, we defined a limb-specific support share based on the positive vertical impulse generated during stance. For each limb, the vertical support impulse was computed as the time integral of vertical ground reaction force exceeding body weight:

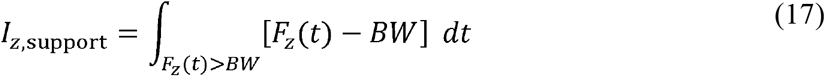

The support share of limb *I* was then defined as:

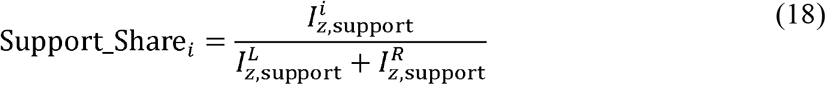

where *L* and *R* denote the left and right limbs, respectively. A value of 0.5 indicates symmetric weight support, whereas deviations reflect asymmetric load-bearing contributions.

This metric quantifies how vertical support is partitioned between limbs during the gait cycle and provides a complementary measure to unloading impulse. By isolating the portion of vertical force exceeding body weight, it captures active load-bearing contributions beyond passive support. In hemiparetic walking, altered support share reflects compensatory redistribution of body weight between limbs and provides a system-level measure of how vertical support constraints shape the mechanical conditions governing step-to-step transition dynamics (Fig. 2).

**Fig. 2:**
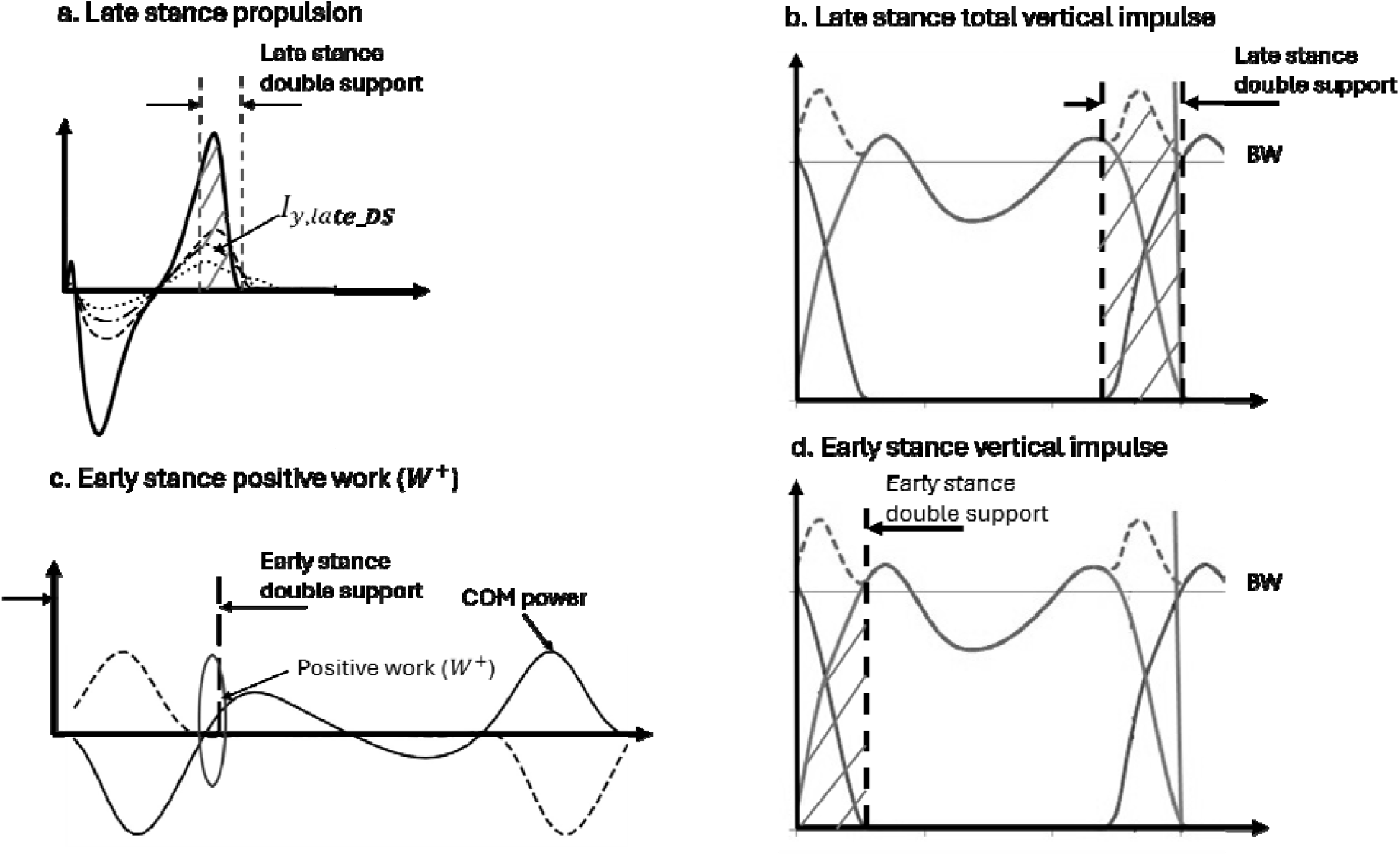
Phase-specific impulse and work reveal feasibility-dependent reorganization of gait mechanics. (a) Late-stance anterior–posterior impulse () reflecting propulsion generation. (b) Late-stance total vertical impulse, representing support during the push-off phase. (c) Early-stance positive COM work, reflecting post-collision recovery, (d) Early-stance vertical impulse associated with collision and load acceptance.

#### Identification of push-off feasibility threshold

The emergence of push-off was evaluated as a function of vertical capacity. A collapse threshold separating push-off absence and presence was estimated using a midpoint criterion between adjacent speed conditions bracketing push-off absence and presence. Uncertainty was quantified using bootstrap resampling across participants.

## Results

### Analytical model identifies dual force thresholds separating support and transition feasibility

The analytical model identified two distinct critical force thresholds governing locomotor feasibility across walking speeds of 0.2–1.2 m·s□^1^ (Fig. 3a). The first threshold corresponded to the minimum force required for vertical load bearing (weight-support threshold: 1.01–1.14 BW), whereas a second, larger threshold defined the minimum force required to generate sufficient mechanical work to redirect the center of mass (COM) during the step-to-step transition (transition threshold: 1.03–1.26 BW). The transition threshold increased with walking speed, consistent with increased energetic demand associated with COM redirection.

**Fig. 3:**
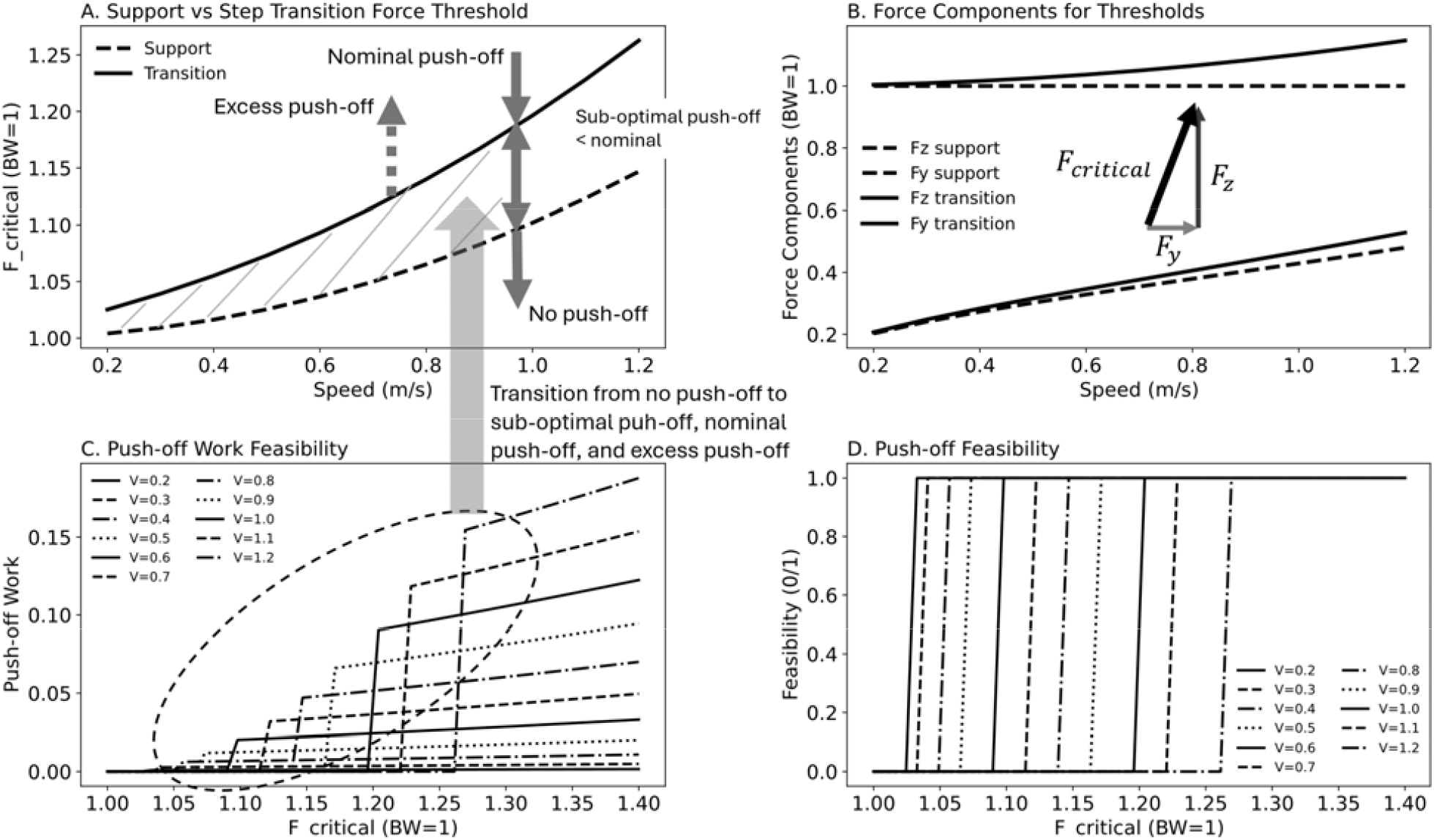
Analytical identification of support and transition thresholds. (a) Force thresholds as a function of walking speed, showing the support feasibility boundary and the higher transition requirement. (b) Decomposition of vertical and anterior– posterior force components contributing to each threshold. (c) Optimized push-off work as a function of available force capacity, showing emergence above the support threshold and progression toward transition-defined work. (d) Binary feasibility map indicating the presence or absence of feasible push-off solutions, demonstrating an abrupt transition at the support boundary.

Decomposition of force components showed that the support threshold was dominated by the vertical component (F_z_ ≈ 1.00 BW), with the anterior–posterior component ranging from 0.20– 0.48 BW across speeds (Fig. 3b). In contrast, the transition threshold required progressively larger anterior–posterior forces as speed increased (F_z_ = 1.00–1.15 BW; F_y_ = 0.21–0.53 BW), separating the mechanical requirements for weight support from those for transition work.

### Optimization reveals abrupt emergence and graded development of push-off

Optimization of late-stance work showed that push-off work increased monotonically with increasing available force capacity once the support threshold was exceeded and approached the analytically predicted transition work as the force bound increased (Fig. 3c).

Binary feasibility analysis demonstrated an abrupt change in the existence of feasible push-off solutions (Fig. 3d). Below the support threshold, no force profile satisfied the vertical impulse constraint and push-off was absent. Above the threshold, feasible solutions emerged abruptly, separating push-off infeasible and push-off capable regimes within the modeled force constraints. Immediately above the support threshold, push-off remained suboptimal and increased progressively toward the analytically predicted transition work as force capacity approached the transition threshold. At fixed speed, optimized transition work closely matched analytical predictions, with efficiency near unity, supporting internal consistency between analytical and optimization formulations.

### Push-off emergence tracks vertical support feasibility in empirical data

Empirically, push-off work depended on system-level vertical feasibility (VCI_sys_) (Fig. 4a–c). At low walking speeds (0.2–0.4 m·s□^1^), system feasibility was near zero or slightly negative (VCI_sys_ = −0.016 to 0.004), and push-off was absent in both limbs. With increasing feasibility at higher speeds (VCI_sys_ = 0.051 at 0.5 m·s□^1^; 0.091 at 0.7 m·s□^1^), push-off emerged in both limbs, with greater magnitude in the unaffected limb (0.064–0.074 J·kg□^1^) than the paretic limb (0.026– 0.051 J·kg□^1^). These findings indicate that the onset of push-off coincides with attainment of minimal vertical support feasibility.

**Fig. 4:**
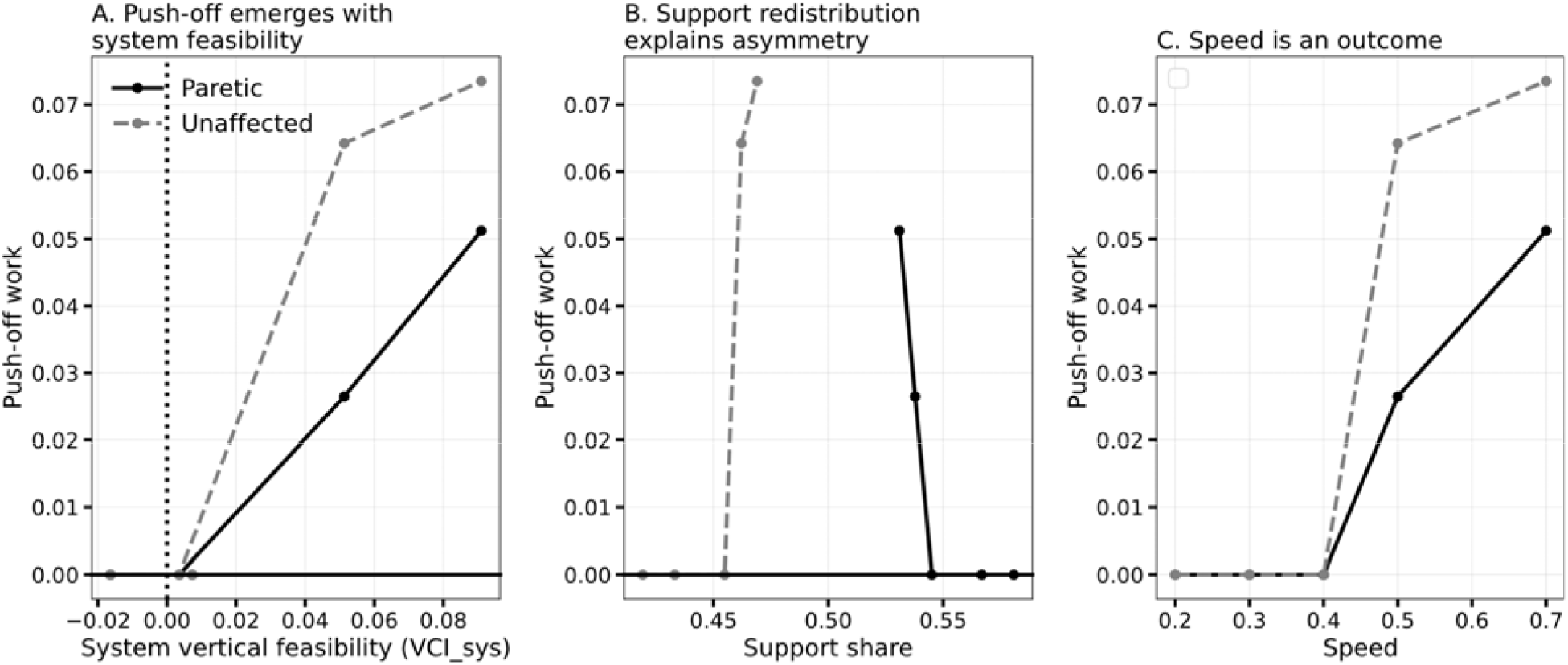
Push-off emergence tracks vertical support feasibility. (a) Push-off work as a function of walking speed for paretic an unaffected limbs. (b) Limb support share during double support. (c) System-level vertical feasibility (VCI). Push-off is absent at low feasibility and emerges once a minimal support threshold is exceeded, with asymmetry between limbs.

Across speeds, the paretic limb contributed a larger fraction of vertical support impulse (support share ≈ 0.53–0.58), whereas the unaffected limb contributed less (≈ 0.42–0.47), and this redistribution persisted after push-off emerged (Fig. 4b). Notably, the limb generating greater push-off work did not correspond to the limb contributing greater vertical support, indicating a dissociation between support and push-off magnitude.

### Unloading capacity tracks the development of push-off

Vertical unloading impulse increased monotonically with walking speed in both limbs, indicating progressive restoration of late-stance unloading capacity (Fig. 5a,c). Push-off work increased concurrently with unloading recovery (Fig. 5a), while unloading remained consistently reduced in the paretic limb.

**Fig. 5:**
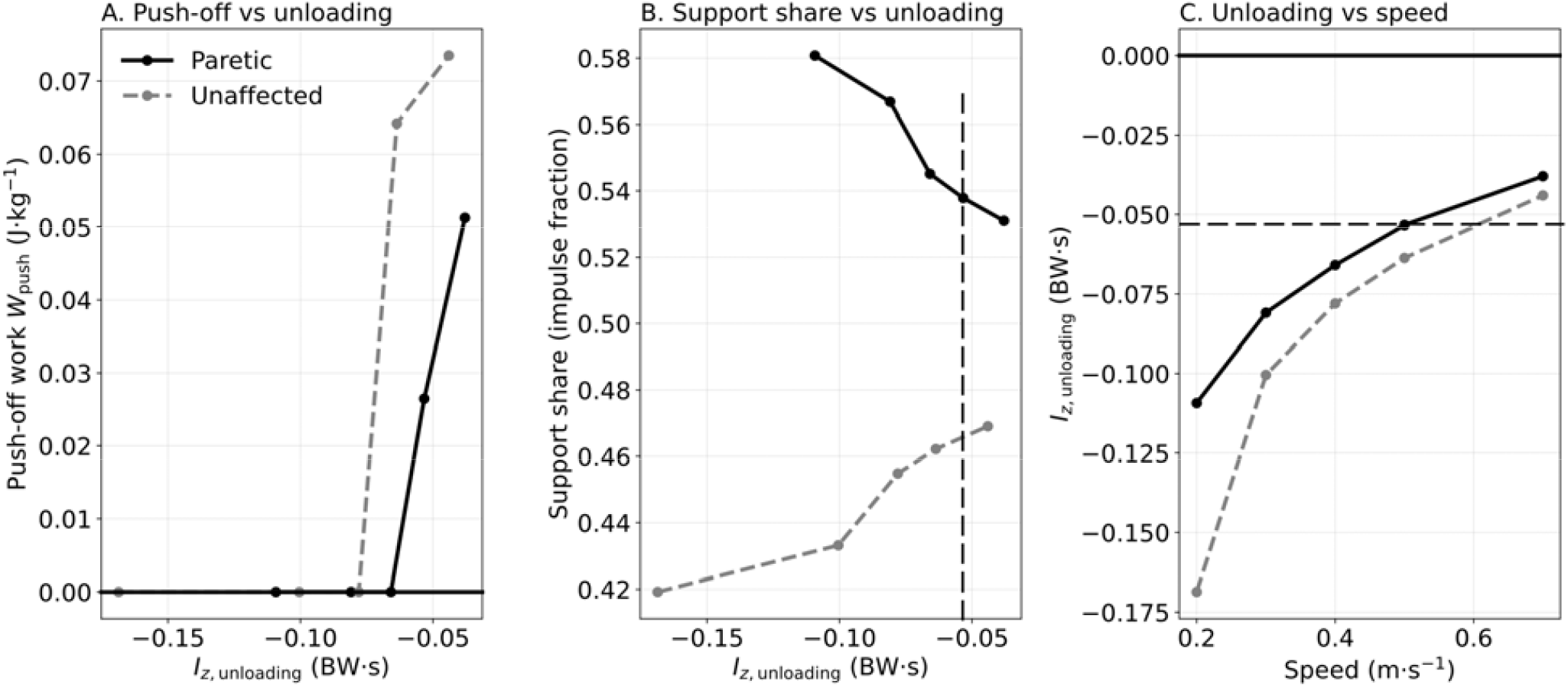
Vertical unloading mediates push-off magnitude. (a) Push-off work versus unloading impulse across speeds. (b) Relationship between unloading and support share. (c) Speed-dependent changes in unloading for paretic and unaffected limbs. Increased unloading is associated with the development and amplification of push-off following its emergence.

Support share varied systematically with unloading capacity (Fig. 5b), such that reduced unloading was associated with compensatory increases in paretic support contribution. The increase in unloading impulse therefore reflects restoration of mechanical conditions associated with the development and amplification of push-off beyond its initial emergence.

### Propulsion capacity can persist without push-off work

The Propulsive Capacity Index (PCI) showed persistent interlimb asymmetry across all walking speeds (Fig 6a–c). The unaffected limb maintained higher PCI values (0.007–0.010), whereas paretic PCI remained near zero (≈0.0004–0.002). Push-off work increased with PCI within each limb (Fig 6b), while PCI remained dissociated from vertical support share (Fig 6c).

**Fig. 6:**
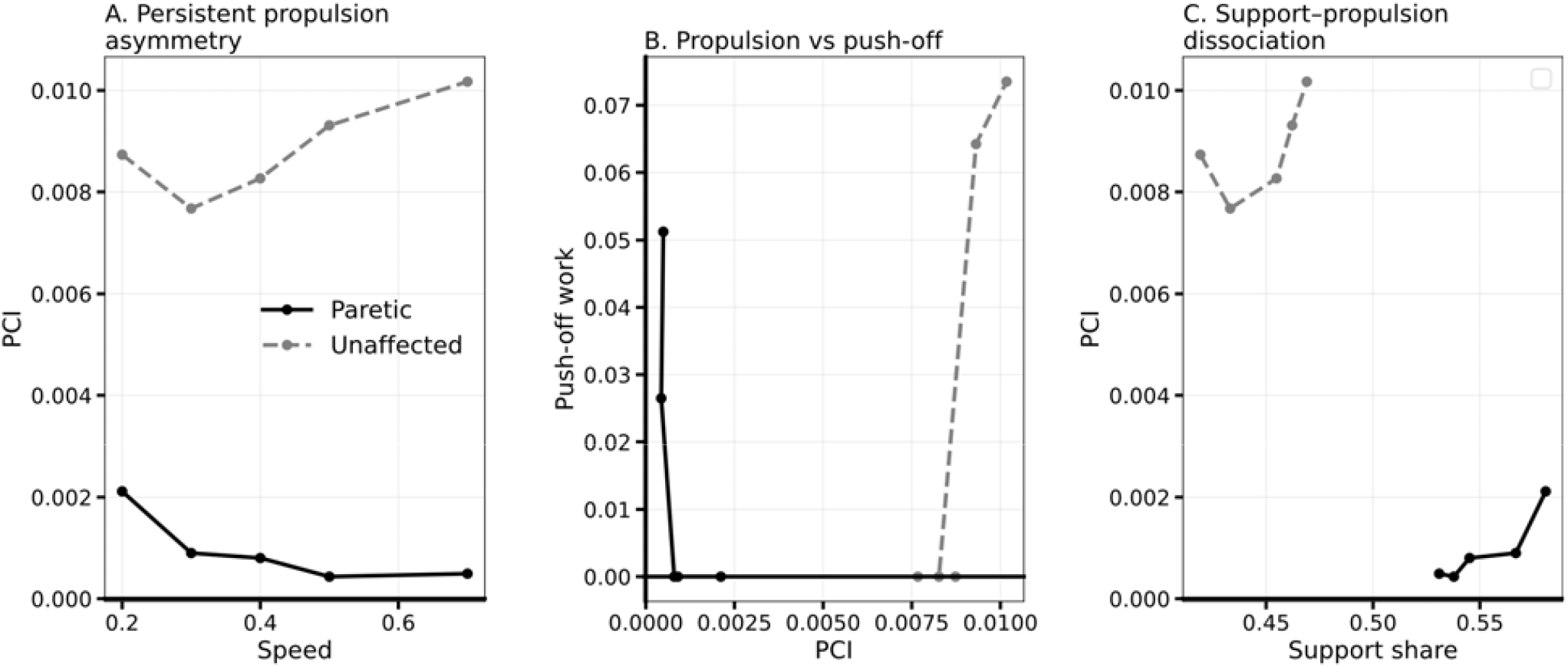
Propulsive capacity persists independently of push-off. (a) Propulsive Capacity Index (PCI) across walking speeds. (b) Relationship between PCI and push-off work. (c) Relationship between PCI and support share. Propulsion varies gradually with speed and does not exhibit a threshold, demonstrating dissociation from push-off emergence.

These results demonstrate that forward-directed propulsive capacity, as captured by PCI, can persist even when push-off work is absent, consistent with push-off being constrained by system-level feasibility rather than propulsion magnitude alone.

### Phase-space analysis reveals a threshold-like transition in gait mechanics

Step-level analysis revealed a structured phase space linking propulsion reserve (PRI), dynamic support demand (RDSF), and collapse probability (Fig 7; Table 1). At low walking speeds (0.2– 0.4 m·s□^1^), paretic collapse probability was elevated (15–60%), whereas collapse was absent at higher speeds once push-off emerged. Unaffected collapse probability increased at intermediate speeds despite preserved support feasibility.

**Table 1:** Graphical indicators used in the stability–capacity diagrams of Fig 7. ‘U’ and ‘P’ represent unaffected and paretic sides respectively.

| Symbol | Representation | Condition | Interpretation |
| --- | --- | --- | --- |
| ● | Filled circle | P limb push trial | Paretic limb step where push-off occurred (no collapse) |
| □ | Hollow square | U limb push trial | Unaffected limb step where push-off occurred (no collapse) |
| × | Cross | Collapse trial (P or U) | Step-to-step transition failure |
| Gray horizontal shading | PRI < threshold | Collapse region | Region where net COM power is insufficient for push-off |
| Gray vertical shading | VCI < threshold | Capacity deficit | Region where vertical support capacity is insufficient |
| Dashed horizontal line | PRI threshold | Push-off onset boundary | Separates collapse from viable push-off |
| Dashed vertical line (Panel A) | RDSF threshold | Redistribution threshold | Boundary in force redistribution dynamics |
| Dashed vertical line (Panel B) | VCI threshold | Mechanical feasibility boundary | Minimum vertical capacity required for push-off |
| Grayscale marker tone | Marker shade | Walking speed encoding | Dark tones indicate slower speeds, and lighter tones indicate faster speeds |

**Fig. 7:**
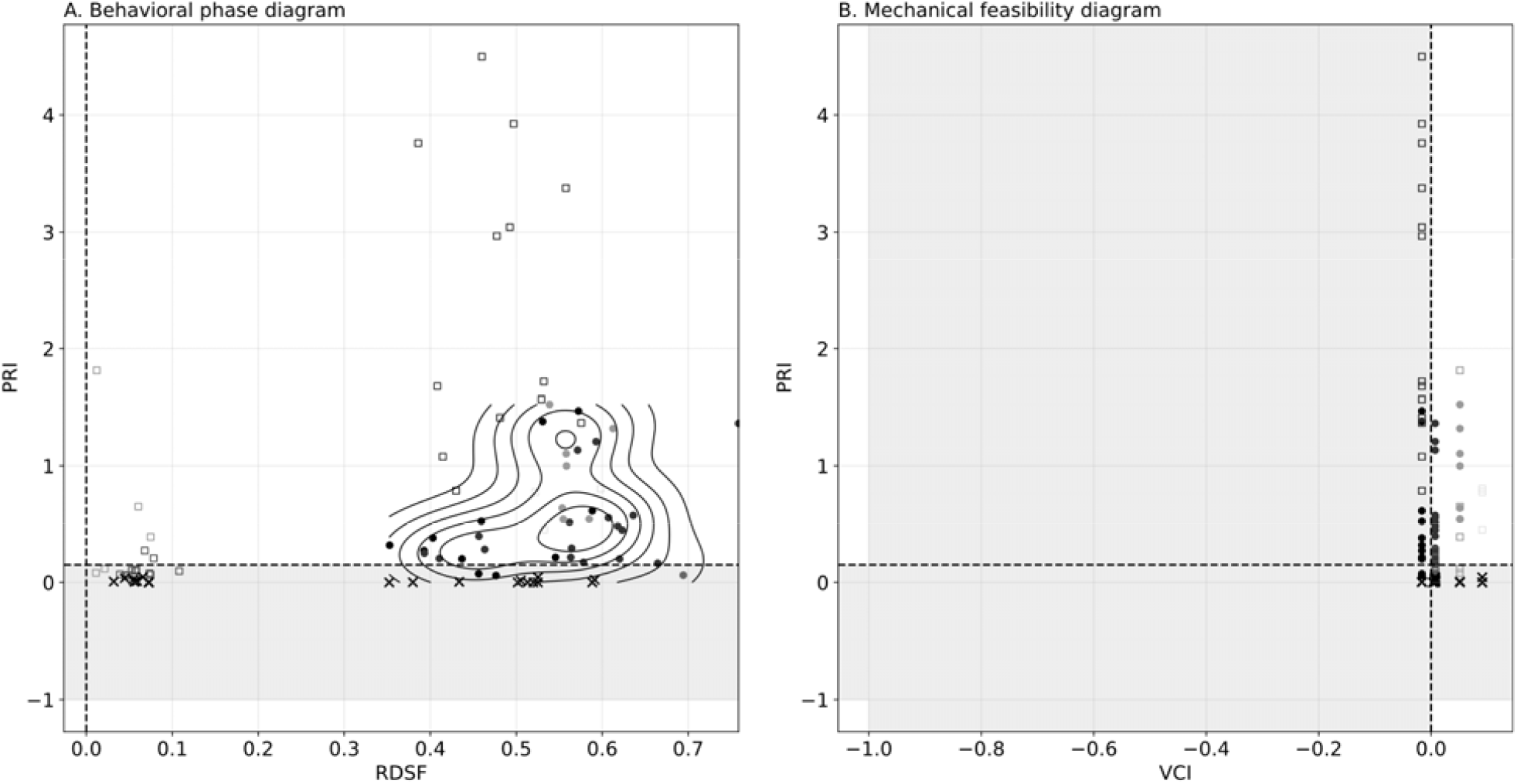
Threshold-like transition in locomotor dynamics. Phase-space representation of propulsion reserve, support demand, and collapse probability. Low-speed walking occupies a support-limited regime characterized by absent push-off and elevated instability, whereas higher speeds transition to a push-off capable regime with stable push-off. The transition occurs over a narrow range and reflects a change in feasible locomotor solutions (Table 1 and Table 2).

PRI increased sharply following push-off emergence, with paretic mean PRI rising from 0.015 at 0.4 m·s□^1^ to 0.95 at 0.5 m·s□^1^ (Fig 7). Collapse events clustered in regions of reduced vertical capacity, whereas push-off dominant steps occupied regions of positive feasibility.

**Table 2:**
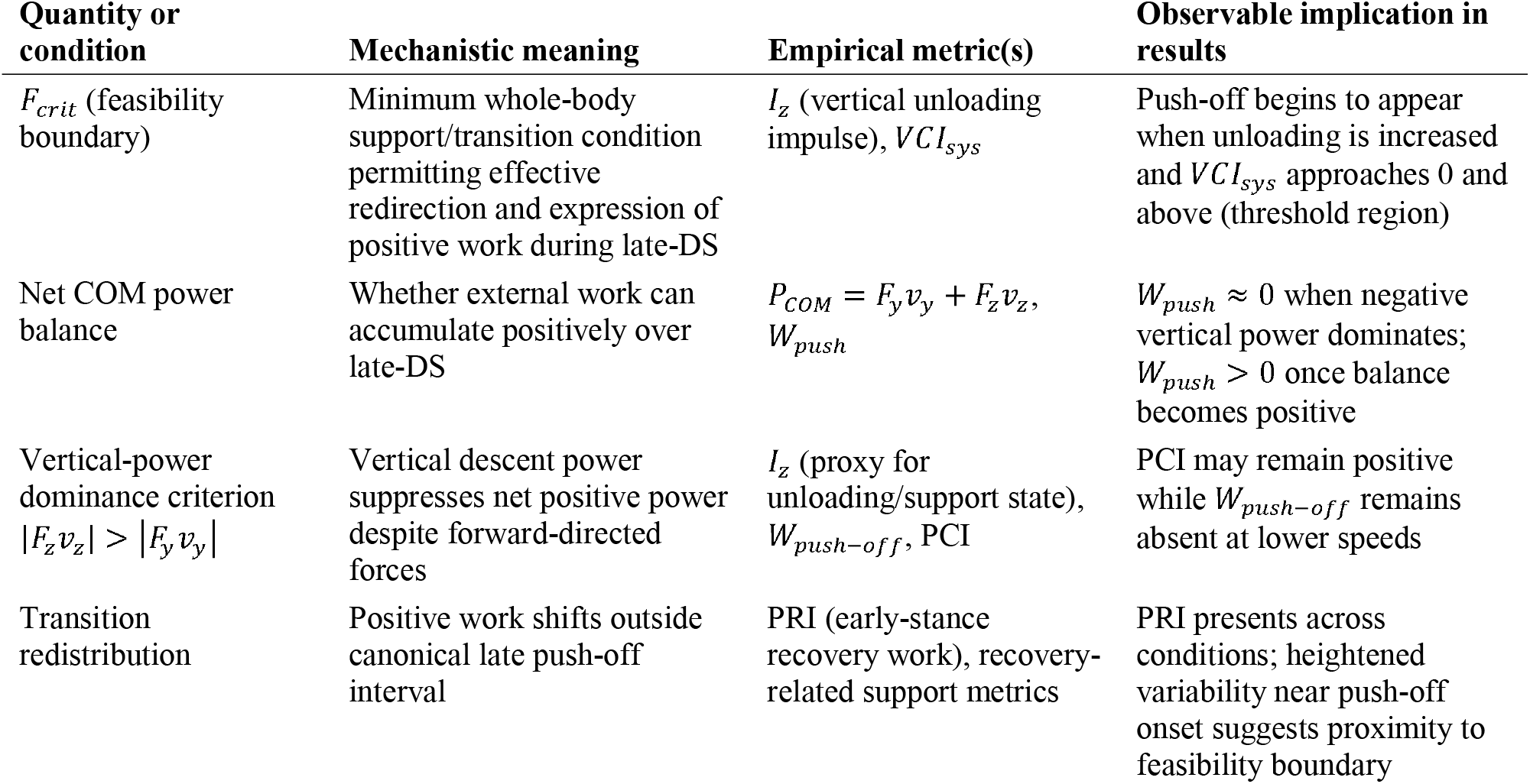
Theoretical–mechanical mapping of support-limited push-off feasibility and its empirical manifestations in COM energetics.

## Discussion

Late-stance push-off is widely treated as the mechanical expression of propulsion capacity in human walking (Huang et al. 2015 Jan 1; Krupenevich et al. 2021). Here we show that this interpretation is incomplete: push-off is not a direct consequence of propulsion generation but a conditional outcome of system-level mechanical feasibility. In the empirical data, forward-directed impulse remains positive even when push-off work is absent, demonstrating that propulsion can persist (Neptune et al. 2001; Awad et al. 2015; Hsiao et al. 2016; Awad et al. 2020) without producing positive center-of-mass (COM) work at late stance. Instead, push-off appears only once system-level vertical feasibility crosses a minimal threshold, indicating that step-to-step transition mechanics (J. Maxwell Donelan et al. 2002; J.Maxwell Donelan et al. 2002; Kuo 2002; Adamczyk and Kuo 2009; Araz et al. 2023) are governed by whole-body constraints rather than propulsion magnitude alone.

This feasibility-based interpretation clarifies why push-off can disappear abruptly rather than degrade continuously with impairment or speed. The analytical framework identifies a hierarchy of constraints: a support feasibility boundary that must be satisfied to maintain vertical load bearing during the transition, and a higher transition requirement associated with COM redirection (Zelik and Adamczyk 2016). This distinction resolves an implicit assumption in classical step-to-step transition theory, which has established the energetic benefit of push-off for reducing collision losses and improving redirection efficiency but typically treats push-off as mechanically available (J. Maxwell Donelan et al. 2002; Kuo 2002; Ruina et al. 2005; Sawicki and Ferris 2009; Hosseini-Yazdi and Bertram 2025 Sept 6). Here, push-off is shown to be conditional: support feasibility governs its emergence, whereas the transition requirement governs its mechanically sufficient or near-optimal expression. In this hierarchy, feasibility precedes efficiency—walking can remain possible without late-stance positive work (push-off), but efficient transition mechanics cannot be expressed until the system enters a push-off capable feasible regime.

A key conceptual outcome of this work is the separation of propulsion from push-off. Propulsion reflects the generation of anterior–posterior force and impulse, whereas push-off requires the conversion of that force into positive COM work during late double support (Kuo et al. 2005). This conversion depends on vertical support conditions (Liu et al. 2008) and force timing, as net COM work reflects the balance of vertical and fore–aft power components. Consequently, propulsion magnitude alone is insufficient to predict push-off. A limb may generate forward-directed impulse (Bowden et al. 2006; Neptune et al. 2009; Roelker et al. 2019; Paskewitz et al. 2025) while remaining unable to accumulate positive work if system-level conditions constrain the late-stance window or cause negative contributions to dominate the net power balance. This distinction resolves the apparent coexistence of persistent propulsion with absent push-off and underscores that reduced push-off should not be attributed solely to diminished propulsive effort.

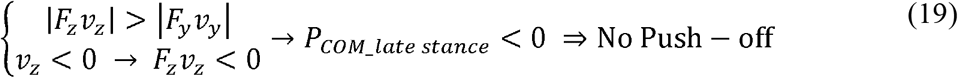

The emergence of push-off is accompanied by a systematic redistribution of mechanical work across the stride, consistent with a shift in gait strategy across feasibility regimes (Hosseini-Yazdi et al. 2026a). When push-off is absent, positive work is redistributed toward early stance, reflecting increased reliance on post-collision recovery. Once push-off becomes feasible, late-stance work increases and early-stance recovery demands diminish, shifting the gait pattern toward transition-driven mechanics characteristic of energetically favorable walking. This redistribution indicates that the system is not simply limited by weakness at low speeds (Patterson et al. 2010) but operates within a regime where late-stance work is mechanically constrained and therefore expressed elsewhere to maintain progression.

Within this framework, vertical unloading capacity provides a mechanistic link between support feasibility and push-off development. Unloading reflects the capacity to reduce vertical force during late stance and transfer load effectively between limbs, creating the conditions under which positive work can accumulate during the transition. The observed association between increased unloading and increased push-off magnitude indicates that recovery of unloading capacity is an intermediate step between the onset of feasibility (push-off becoming possible) and the progressive strengthening of push-off (approaching transition-defined work). Persistent reductions in paretic unloading align with reduced paretic push-off, reinforcing that the limiting factor is not propulsion intent, but the mechanical context required to express propulsion as positive transition work.

Taken together, the analytical and empirical findings indicate a threshold-driven transition between qualitatively distinct locomotor regimes. Below the support feasibility boundary, walking is characterized by reduced vertical capacity, absence of push-off, and increased collapse probability; above it, push-off emerges abruptly and transition mechanics stabilize. This transition occurs over a narrow range of walking speeds and is not explained by gradual changes in propulsion alone. Rather, it reflects a change in the set of mechanically feasible solutions governing COM dynamics, consistent with a bifurcation in locomotor behavior defined by feasibility constraints.

Because these constraints arise from whole-body mechanics rather than pathology-specific deficits (Peterson et al. 2010; Awad et al. 2015; Awad et al. 2015; Lewek et al. 2018; Roelker et al. 2019; Alam et al. 2022; Paskewitz et al. 2025; Paskewitz et al. 2025), the identified feasibility structure is expected to govern locomotion more broadly, including unimpaired walking. This generalization elevates push-off from a symptom of impairment to a manifestation of fundamental mechanical constraints (Bertram and Ruina 2001; Bertram 2005).

These findings have direct implications for rehabilitation and assistive control. If push-off is a conditional outcome of feasibility, interventions targeting propulsion (Bowden et al. 2006; Lewek et al. 2018; Roelker et al. 2019) in isolation may be ineffective when vertical support capacity remains limiting. Restoring support feasibility—through strengthening (Choi et al. 2020), coordination training, or assistance (Roerdink et al. 2007)—may be a prerequisite for enabling push-off to emerge, after which propulsion-focused interventions can improve symmetry and efficiency. For assistive technologies (Sawicki and Ferris 2009), these results motivate control strategies that prioritize vertical support and unloading capacity before augmenting propulsion, and suggest that ground reaction force–derived feasibility metrics may provide a basis for detecting regime transitions and adapting assistance in real time (Fig. 8, Table 2).

**Fig. 8:**
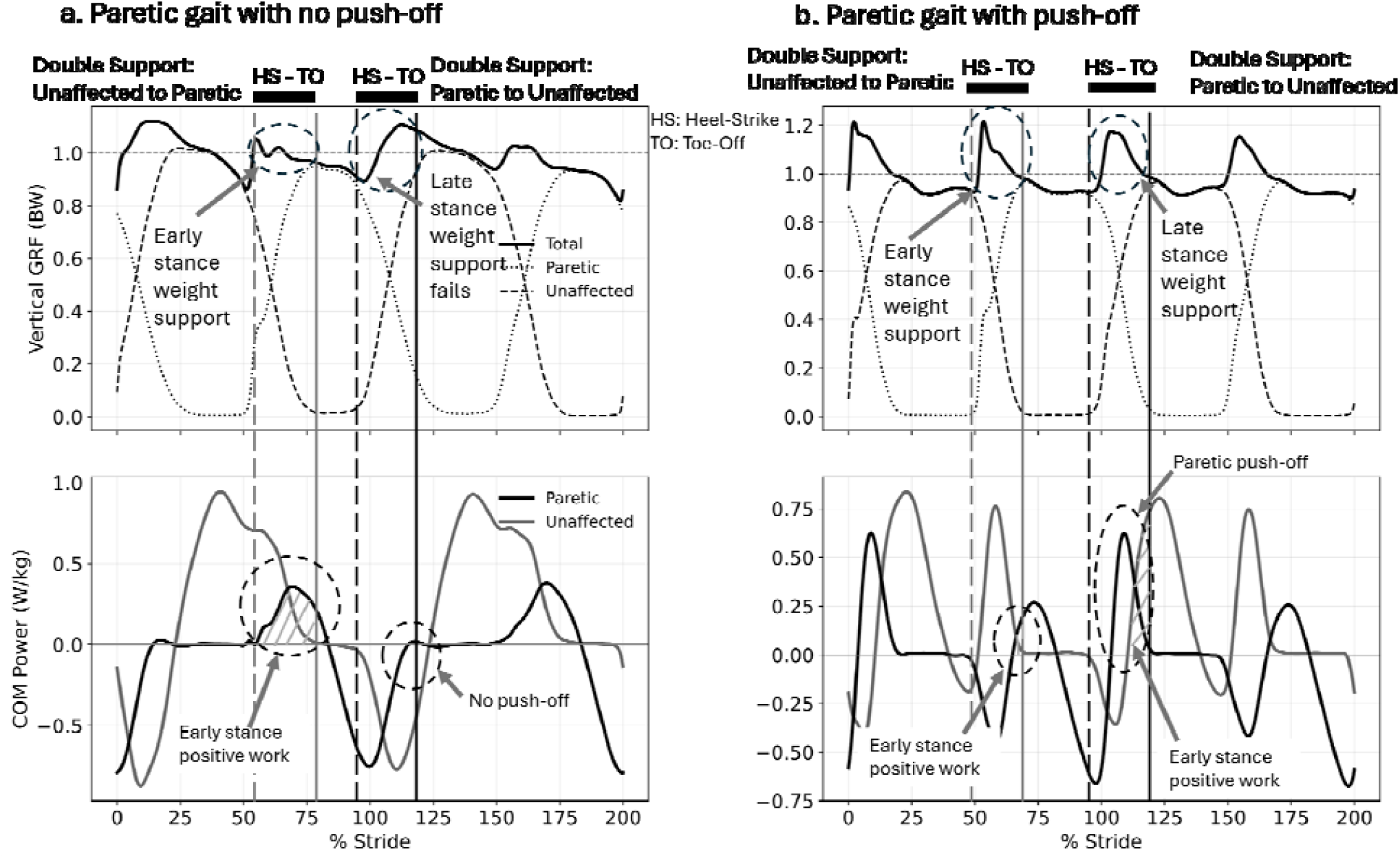
Representative vertical ground reaction force (Fz) and center-of-mass (COM) power illustrate the role of support feasibility in push-off generation. (a) Example condition with negative system-level vertical feasibility. Although vertical ground reaction force is present, it is insufficient relative to the feasibility requirement, and no late-stance positive COM power is observed, indicating absence of push-off. (b) Example condition with positive vertical feasibility. Under these conditions, sufficient support capacity is achieved, and late-stance positive COM power emerges, indicating the presence of push-off (Table 2).

Several limitations should be acknowledged. The analytical model is intentionally reduced-order and assumes force alignment with the stance limb, without explicitly representing joint-level actuation or neuromuscular control (Farris et al. 2015; Browne and Franz 2017; Browne and Franz 2018; Delabastita et al. 2021). While this abstraction isolates fundamental mechanical constraints, future work should examine how these feasibility boundaries interact with muscle coordination, joint-level impairments, and altered timing. In addition, the present cohort reflects subacute post-stroke walking at relatively low speeds; extending this framework to higher speeds, overground conditions (Lee and Hidler 2008), and diverse populations will be important for establishing generality and translational scope.

In summary, push-off is not a direct consequence of propulsion generation but a feasibility-limited outcome governed by vertical support constraints. Support feasibility enables its emergence, whereas the transition requirement governs its mechanically sufficient expression. By establishing push-off as a feasibility-limited phenomenon, this work reframes its role in locomotion, resolves its dissociation from propulsion, and provides a unified mechanical basis for understanding gait impairment, recovery, and control.

## Data Availability

GitHub at: salehhosseini/Hemiparetic-dataset: Hemi Paretic Walking Dataset

## Ethical approval

Subjects provided written informed consent (Ethics ID REB21-1576)

## Author contributions

S-S Hosseini-Yazdi – Conception of study, analysis, writing and editing; K Fitzsimons – Empirical data collection and analysis, writing and editing; JEA Bertram – Planning of study, resourcing, writing and editing.

## Data availability

The datasets used in this study are available in the author’s public repository on GitHub at: <u>salehhosseini/Hemiparetic-dataset: Hemi Paretic Walking Dataset</u>

## References

Adamczyk PG, Kuo AD. 2009. Redirection of center-of-mass velocity during the step-to-step transition of human walking. J Exp Biol. 212(Pt 16):2668–2678. 10.1242/jeb.027581

Alam Z et al. 2022. Timing of propulsion-related biomechanical variables is impaired in individuals with post-stroke hemiparesis. Gait & Posture. 96:275–278. 10.1016/j.gaitpost.2022.05.022

Alexander RM. 1992. Simple models of walking and jumping. Human Movement Science. 11(1– 2):3–9. 10.1016/0167-9457(92)90045-D

Alexander RMcN. 1995. Simple Models of Human Movement. Applied Mechanics Reviews. 48(8):461–470. 10.1115/1.3005107

Allen JL, Kautz SA, Neptune RR. 2011. Step length asymmetry is representative of compensatory mechanisms used in post-stroke hemiparetic walking. Gait Posture. 33(4):538– 543. 10.1016/j.gaitpost.2011.01.004

Anderson FC, Pandy MG. 2003. Individual muscle contributions to support in normal walking. Gait Posture. 17(2):159–169. 10.1016/s0966-6362(02)00073-5

Araz M et al. 2023. Muscle preflex response to perturbations in locomotion: In vitro experiments and simulations with realistic boundary conditions. Front Bioeng Biotechnol. 11:1150170. 10.3389/fbioe.2023.1150170

Awad LN et al. 2020. These legs were made for propulsion: advancing the diagnosis and treatment of post-stroke propulsion deficits. J NeuroEngineering Rehabil. 17(1):139. 10.1186/s12984-020-00747-6

Awad LN, Binder-Macleod SA, Pohlig RT, Reisman DS. 2015. Paretic Propulsion and Trailing Limb Angle Are Key Determinants of Long-Distance Walking Function After Stroke. Neurorehabil Neural Repair. 29(6):499–508. 10.1177/1545968314554625

Balasubramanian CK, Bowden MG, Neptune RR, Kautz SA. 2007. Relationship between step length asymmetry and walking performance in subjects with chronic hemiparesis. Arch Phys Med Rehabil. 88(1):43–49. 10.1016/j.apmr.2006.10.004

Beaman CB, Peterson CL, Neptune RR, Kautz SA. 2010. Differences in self-selected and fastest-comfortable walking in post-stroke hemiparetic persons. Gait Posture. 31(3):311–316. 10.1016/j.gaitpost.2009.11.011

Bertram JEA. 2005. Constrained optimization in human walking: cost minimization and gait plasticity. Journal of Experimental Biology. 208(6):979–991. 10.1242/jeb.01498

Bertram JEA, Ruina A. 2001. Multiple Walking Speed–frequency Relations are Predicted by Constrained Optimization. Journal of Theoretical Biology. 209(4):445–453. 10.1006/jtbi.2001.2279

Bowden MG, Balasubramanian CK, Neptune RR, Kautz SA. 2006. Anterior-Posterior Ground Reaction Forces as a Measure of Paretic Leg Contribution in Hemiparetic Walking. Stroke. 37(3):872–876. 10.1161/01.STR.0000204063.75779.8d

Browne MG, Franz JR. 2017. The independent effects of speed and propulsive force on joint power generation in walking. Journal of Biomechanics. 55:48–55. 10.1016/j.jbiomech.2017.02.011

Browne MG, Franz JR. 2018. More push from your push-off: Joint-level modifications to modulate propulsive forces in old age Grabowski A, editor. PLoS ONE. 13(8):e0201407. 10.1371/journal.pone.0201407

Cavagna GA, Heglund NC, Taylor CR. 1977. Mechanical work in terrestrial locomotion: two basic mechanisms for minimizing energy expenditure. Am J Physiol. 233(5):R243–261. 10.1152/ajpregu.1977.233.5.R243

Choi EP et al. 2020. Changes in Lower Limb Muscle Activation and Degree of Weight Support according to Types of Cane-Supported Gait in Hemiparetic Stroke Patients. Biomed Res Int. 2020:9127610. 10.1155/2020/9127610

Dar G, Saposhnik A, Finestone AS, Ayalon M. 2023. The Effect of Load Carrying on Gait Kinetic and Kinematic Variables in Soldiers with Patellofemoral Pain Syndrome. Applied Sciences. 13(4):2264. 10.3390/app13042264

Darici O, Temeltas H, Kuo AD. 2018. Optimal regulation of bipedal walking speed despite an unexpected bump in the road Haddad JM, editor. PLoS ONE. 13(9):e0204205. 10.1371/journal.pone.0204205

Delabastita T et al. 2021. Distal*to*proximal joint mechanics redistribution is a main contributor to reduced walking economy in older adults. Scandinavian Med Sci Sports. 31(5):1036–1047. 10.1111/sms.13929

Donelan J. Maxwell, Kram R, Kuo AD. 2002. Simultaneous positive and negative external mechanical work in human walking. Journal of Biomechanics. 35(1):117–124. 10.1016/S0021-9290(01)00169-5

Donelan J. Maxwell, Kram R, Kuo AD. 2002. Mechanical work for step-to-step transitions is a major determinant of the metabolic cost of human walking. J Exp Biol. 205(Pt 23):3717–3727. 10.1242/jeb.205.23.3717

Farris DJ, Hampton A, Lewek MD, Sawicki GS. 2015. Revisiting the mechanics and energetics of walking in individuals with chronic hemiparesis following stroke: from individual limbs to lower limb joints. J NeuroEngineering Rehabil. 12(1):24. 10.1186/s12984-015-0012-x

Hall KD et al. 2011. Quantification of the effect of energy imbalance on bodyweight. The Lancet. 378(9793):826–837. 10.1016/S0140-6736(11)60812-X

Hansen AH, Childress DS, Meier MR. 2002. A simple method for determination of gait events. J Biomech. 35(1):135–138. 10.1016/s0021-9290(01)00174-9

Hosseini-Yazdi S-S, Bertram JE. 2025. The consequence of uneven walking transitory modulation strategies: A simulation-based approach. J Theor Biol. 614:112234. 10.1016/j.jtbi.2025.112234

Hosseini-Yazdi S-S, Bertram JEA. 2025. Center of mass work analysis predicts preferred walking speeds for varying walking conditions. Journal of Biomechanics. 185:112682. 10.1016/j.jbiomech.2025.112682

Hosseini-Yazdi S-S, Bertram JEA. 2025. Optimum Push-off for Uneven Walking Based on the Just-in-Time Strategy: Walking with Interrupted Push-Off is Mechanically Costly. Journal of Biomechanical Engineering. 1–20. 10.1115/1.4069666

Hosseini-Yazdi S-S, Fitzsimons K, Bertram JE. 2026a. Mechanistic Reorganization of Step Work in Hemiparetic Walking: Modeling and Center-of-Mass Power/Work Analysis. [accessed 2026 Mar 28]. http://medrxiv.org/lookup/doi/10.64898/2026.03.11.26348174. 10.64898/2026.03.11.26348174

Hosseini-Yazdi S-S, Fitzsimons K, Bertram JE. 2026b. Center-of-Mass Work Organization Supplements Walking Speed: a Biomechanical Characterization of Hemiparetic Gait. [accessed 2026 Mar 28]. http://medrxiv.org/lookup/doi/10.64898/2026.03.12.26348298. 10.64898/2026.03.12.26348298

Hosseini-Yazdi S-S, Kuo AD. 2025. The energetic cost of human walking as a function of uneven terrain amplitude. Journal of Experimental Biology. jeb.249840. 10.1242/jeb.249840

Hsiao H et al. 2016. Contribution of Paretic and Nonparetic Limb Peak Propulsive Forces to Changes in Walking Speed in Individuals Poststroke. Neurorehabil Neural Repair. 30(8):743– 752. 10.1177/1545968315624780

Huang TP, Shorter KA, Adamczyk PG, Kuo AD. 2015. Mechanical and energetic consequences of reduced ankle plantarflexion in human walking. Journal of Experimental Biology. [published online ahead of print] [accessed 2025 July 22]. https://journals.biologists.com/jeb/article/doi/10.1242/jeb.113910/258238/Mechanical-and-energetic-consequences-of-reduced. 10.1242/jeb.113910

Jung D-E, Kim K. 2015. The effects of ankle loads on balance ability during one-leg stance. J Phys Ther Sci. 27(5):1527–1528. 10.1589/jpts.27.1527

Krupenevich RL et al. 2021. Effects of age and locomotor demand on foot mechanics during walking. Journal of Biomechanics. 123:110499. 10.1016/j.jbiomech.2021.110499

Kuo AD. 2001. A simple model of bipedal walking predicts the preferred speed-step length relationship. J Biomech Eng. 123(3):264–269. 10.1115/1.1372322

Kuo AD. 2002. Energetics of actively powered locomotion using the simplest walking model. J Biomech Eng. 124(1):113–120. 10.1115/1.1427703

Kuo AD, Donelan JM, Ruina A. 2005. Energetic Consequences of Walking Like an Inverted Pendulum: Step-to-Step Transitions: Exercise and Sport Sciences Reviews. 33(2):88–97. 10.1097/00003677-200504000-00006

Lee SJ, Hidler J. 2008. Biomechanics of overground vs. treadmill walking in healthy individuals. Journal of Applied Physiology. 104(3):747–755. 10.1152/japplphysiol.01380.2006

Lewek MD, Raiti C, Doty A. 2018. The Presence of a Paretic Propulsion Reserve During Gait in Individuals Following Stroke. Neurorehabil Neural Repair. 32(12):1011–1019. 10.1177/1545968318809920

Liu MQ, Anderson FC, Schwartz MH, Delp SL. 2008. Muscle contributions to support and progression over a range of walking speeds. Journal of Biomechanics. 41(15):3243–3252. 10.1016/j.jbiomech.2008.07.031

Neptune RR, Clark DJ, Kautz SA. 2009. Modular control of human walking: A simulation study. Journal of Biomechanics. 42(9):1282–1287. 10.1016/j.jbiomech.2009.03.009

Neptune RR, Kautz SA, Zajac FE. 2001. Contributions of the individual ankle plantar flexors to support, forward progression and swing initiation during walking. Journal of Biomechanics. 34(11):1387–1398. 10.1016/s0021-9290(01)00105-1

Olney SJ, Richards C. 1996. Hemiparetic gait following stroke. Part I: Characteristics. Gait & Posture. 4(2):136–148. 10.1016/0966-6362(96)01063-6

Paskewitz J, Fei J, Wang R, Awad LN. 2025. Classifying Post-Stroke Gait Propulsion Impairment Beyond Walking Speed: A Clinically Feasible Approach Using the Functional Gait Assessment. Applied Sciences. 16(1):134. 10.3390/app16010134

Patterson KK et al. 2010. Evaluation of gait symmetry after stroke: A comparison of current methods and recommendations for standardization. Gait & Posture. 31(2):241–246. https://doi.org/16/j.gaitpost.2009.10.014

Peterson CL, Hall AL, Kautz SA, Neptune RR. 2010. Pre-swing deficits in forward propulsion, swing initiation and power generation by individual muscles during hemiparetic walking. J Biomech. 43(12):2348–2355. 10.1016/j.jbiomech.2010.04.027

Roelker SA, Bowden MG, Kautz SA, Neptune RR. 2019. Paretic propulsion as a measure of walking performance and functional motor recovery post-stroke: A review. Gait & Posture. 68:6– 14. 10.1016/j.gaitpost.2018.10.027

Roerdink M et al. 2007. Gait coordination after stroke: benefits of acoustically paced treadmill walking. Phys Ther. 87(8):1009–1022. 10.2522/ptj.20050394

Ruina A, Bertram JEA, Srinivasan M. 2005. A collisional model of the energetic cost of support work qualitatively explains leg sequencing in walking and galloping, pseudo-elastic leg behavior in running and the walk-to-run transition. Journal of Theoretical Biology. 237(2):170–192. 10.1016/j.jtbi.2005.04.004

Sawicki GS, Ferris DP. 2009. Powered ankle exoskeletons reveal the metabolic cost of plantar flexor mechanical work during walking with longer steps at constant step frequency. Journal of Experimental Biology. 212(1):21–31. 10.1242/jeb.017269

Sheffler LR, Chae J. 2015. Hemiparetic Gait. Physical Medicine and Rehabilitation Clinics of North America. 26(4):611–623. 10.1016/j.pmr.2015.06.006

Soo CH, Donelan JM. 2010. Mechanics and energetics of step-to-step transitions isolated from human walking. Journal of Experimental Biology. 213(24):4265–4271. 10.1242/jeb.044214

White H, Uhl TL, Augsburger S, Tylkowski C. 2007. Reliability of the three-dimensional pendulum test for able-bodied children and children diagnosed with cerebral palsy. Gait & Posture. 26(1):97–105. 10.1016/j.gaitpost.2006.07.012

Zelik KE, Adamczyk PG. 2016. A unified perspective on ankle push-off in human walking. Journal of Experimental Biology. 219(23):3676–3683. 10.1242/jeb.140376

Zeni JA, Richards JG, Higginson JS. 2008. Two simple methods for determining gait events during treadmill and overground walking using kinematic data. Gait Posture. 27(4):710–714. 10.1016/j.gaitpost.2007.07.007

